# Development and evaluation of a multivariate prediction model for diagnosing asthma in patients with clinically suspected asthma using capnography

**DOI:** 10.1101/2025.11.25.25340952

**Authors:** Henry Broomfield, Leeran Talker, Rui Hen Lim, Joseph Massingham, Gabriel Lambert, Helen Ashdown, Gail Hayward, Daniel Neville, Thomas Brown, Laura Wiffen, Anoop Chauhan, Ameera X. Patel

## Abstract

**Background:** Diagnosis of asthma in primary care is challenged by a multistep pathway with variable adherence leading to significant misdiagnosis, late diagnosis and poor patient outcomes. This is driven by a lack of rapid, accurate, easy-to-use diagnostic tests that can reliably diagnose asthma with both high sensitivity and specificity. The objective of this work was to develop and evaluate a multivariate machine learning classifier for asthma diagnosis. The classifier was built using interpretable data processing and machine learning techniques applied to 75-second tidal breathing CO_2_ recordings captured on TidalSense’s N-Tidal^®^ hand-held capnometer. The target population comprises patients with clinically suspected asthma who have no contraindications to performing capnometry.

**Methods:** Capnograms were collected from 138 asthmatic and 132 non-asthmatic participants (including healthy volunteers, and those with chronic obstructive pulmonary disease (COPD), heart failure, and other cardiorespiratory conditions) recruited from both primary and secondary care. Each high-resolution CO_2_ recording was transformed into 82 features (using the N-Tidal^®^ Diagnose 1 v1.0 software) that characterise the constituent breathing cycles. A logistic regression model was trained on these features and performance metrics generated from an unseen test set of 64 participants. Model performance was evaluated using discrimination, measured by the area under the receiver operating characteristic curve (AUROC), as well as clinically relevant predictive accuracy metrics, including positive predictive value (PPV) and negative predictive value (NPV). This was repeated 20 times with different training and testing participants for additional statistical robustness; the average and variability of these metrics were recorded.

**Results:** The classification model achieved an AUROC of 0.91 ± 0.03%, sensitivity of 83 ± 4%, specificity of 85 ± 6%, positive predictive value (PPV) of 87 ± 4%, and negative predictive value (NPV) of 81 ± 4% in detecting asthma from a single breath recording. The model demonstrated diagnostic stability, with 95.8% of each participant’s recordings over the course of data collection being classified correctly on average. No model bias was observed with regards to sex, but performance did improve with age, possibly reflecting increasing severity of disease with age.

**Conclusion:** This study introduces a highly accurate and interpretable multivariate diagnostic model capable of classifying asthma from a single breath recorded using the N-Tidal^®^ Handset. It achieves high sensitivity and specificity compared with current methods, such as spirometry, and could enable point-of-care diagnosis in patients suspected of having asthma.

## Introduction

Asthma is a major global public health challenge as a condition with both high prevalence and high incidence (Sinyor et al. 2023). It is characterised by airway inflammation and hyper-responsiveness, primarily of the conducting zone of the bronchial tree. Asthma affects people of all ages and is the most common long-term condition among children in the UK (NHS 2021). In 2019 asthma was estimated to affect 262 million people worldwide – resulting in over 450,000 deaths (WHO 2023). In the UK alone, asthma carries an annual fiscal burden to the NHS of £1.1 billion (Mukherjee et al. 2016).

Early diagnosis and treatment of asthma are important to reduce symptoms, prevent exacerbations and mortality, and improve overall quality of life (Kupczyk et al. 2010). However, its timely and accurate diagnosis is hindered by the absence of a single test that can accurately diagnose asthma (NICE 2023). This gives rise to clinical diagnostic pathways that are often poorly followed and involve multiple tests that have limited accessibility in primary care. As a result, there is a high prevalence of empirical diagnosis (Pavord et al. 2018), which leads to significant misdiagnosis and poor outcomes for patients.

Spirometry is a universal component of asthma diagnostic pathways, quantifying airway obstruction by measuring the volume and flow of exhaled air. A hallmark feature of asthma is the reversibility of airway obstruction, which is assessed using bronchodilator reversibility (BDR) testing. BDR involves performing spirometry before and after bronchodilator administration and evaluating the change in lung function to determine the degree of reversible airway obstruction. However, spirometry, and by extension BDR testing, is technique dependent and operationally challenging to deliver, requiring accredited professionals and those competent in interpretation to deliver an accurate diagnosis. Furthermore, its accuracy depends on patients being medication naïve, meaning BDR testing has historically had limited uptake in primary care. In the UK, concerns about aerosol generation associated with spirometry or cough associated with the procedure during the COVID-19 pandemic led to a halting of spirometry services in primary care many of which have still not restarted (NHS 2024). Marked regional variations now exist in access to spirometry with concerns that this particularly impacts areas of high socio-economic deprivation (Howard 2023). These factors, combined with the low sensitivity of spirometry-based BDR testing when interpreted according to most diagnostic guidelines (Simpson et al. 2024), may have contributed to an underdiagnosis rate for asthma that is estimated to be between 20% and 70% (Aaron et al. 2018).

As a result of this, recent guideline changes in the UK now recommend alternative testing with blood eosinophil measurements and fractional inhaled nitric oxide (FeNO) (NICE 2024a) as first line diagnostic tests for asthma. These may be easier to deliver in primary care but are challenged by poor sensitivity (32% and 53% respectively (NICE 2024c)) and can only diagnose patients with type-2 airway inflammation (i.e. eosinophilic asthma). This reserves spirometry testing for those who cannot be definitively diagnosed with either FeNO or blood eosinophil count, with a pathway ending up in bronchial hyperresponsiveness challenge if spirometry and peak expiratory flow testing is inconclusive. It is estimated that 62% of patients (NICE 2024c) would require bronchial challenge (to which access is limited, even in secondary care, due to cost and the low availability of methacholine (NICE 2024d)) for a definitive diagnosis. Consequently, there is a significant need for new, accurate, easy-to-deliver, asthma diagnostic testing in primary care that can be delivered reliably and would ideally maintain diagnostic accuracy for those in whom empirical treatment has already been started.

Capnometry, the measurement of respired carbon dioxide CO_2_ concentration, is a widely used technique in critical care. CO_2_ measurements are routinely used to monitor respiratory rate and end-tidal CO_2_ concentration to detect episodes of hypoventilation, ventilatory or circulatory failure, and malfunction or malposition of endotracheal tubes. While it has been acknowledged that capnography can provide information on airway obstruction (Jaffe 2017; Pertzov et al. 2021), use in respiratory diagnosis has been limited due to the lack of fast-response CO_2_ sensing capabilities that provide a high signal-to-noise ratio while maintaining fine resolution in the information-rich regions of the signal. The geometric properties of capnograms have been used to classify both COPD and asthma patients as distinct from healthy individuals (Mieloszyk et al. 2014; Singh et al. 2018). More recently, Talker et al. (2023) provided the first demonstration of accurate diagnostic machine learning classification of COPD from common differential diagnoses for COPD, using the N-Tidal^®^ time-based capnometer (N-Tidal^®^ Handset). CO_2_-waveform features in the region of gas transition from large airways to alveolar gas demonstrated good correlation with FEV_1_% predicted across all COPD GOLD stages. The ability to grade COPD severity using volumetric capnography and deep learning has also been explored (Mou et al. 2024).

This manuscript aims to develop and evaluate the first proof-of-concept diagnostic classification of asthma with time-based capnometry using the N-Tidal^®^ Handset, which has potential to enable rapid point-of-care testing in asthma diagnostic pathways. The presented model is intended for use by healthcare professionals to support the diagnosis of patients with clinically suspected asthma.

## Methods

Updated Standards for Reporting of Diagnostic Accuracy Studies (STARD) (Cohen et al. 2016) and Transparent Reporting of Multivariable Prediction Models for Individual Prognosis or Diagnosis with Artificial Intelligence (TRIPOD AI) (Collins et al. 2024) have been utilised to ensure transparency and thoroughness of reporting.

### Participants

The data used in this research was compiled from four longitudinal observational studies (ABRS, CBRS, CBRS2, GBRS), conducted between February 2016 to January 2022. Each study recruited participants with different pre-existing cardiorespiratory conditions, including those that are common differential diagnoses for asthma. These studies included participants with asthma, breathing pattern disorder (BPD), COPD, heart failure (CHF), motor neuron disease (MND), pneumonia, and those without any medical conditions (healthy). In addition to these four studies, capnography data was collected using the N-Tidal^®^ Handset from 28 volunteers (healthy controls) without any respiratory disease between December 2015 and December 2022. These volunteers provided written informed consent and were screened by a medical doctor to ensure they did not have any cardiorespiratory disease. Together, participants from the above studies formed a heterogeneous dataset that was designed to be representative of the population that could be referred for asthma diagnostic testing.

In participants with asthma, diagnoses were made according to accepted guidelines including NICE, BTS/SIGN or GINA (see supplementary material for specific diagnostic criteria in each study). Ethical approval was obtained from the South Central - Berkshire - Research Ethics Committee (REC) for ABRS and GBRS, the Yorkshire and the Humber REC for CBRS and the West Midlands Solihull REC for CBRS2. All participants across the clinical studies gave informed consent, and their data was handled according to all applicable data protection legislations, including EU/UK General Data Protection Regulation. Details of all studies, including objectives and eligibility criteria, have been provided in the supplementary material.

### Procedures

Participants were provided a training session on the appropriate use of the N-Tidal^®^ Handset from a trained instructor. Participants were instructed to perform tidal breathing at rest through the mouth for 75 seconds. This produced a 50 Hz breath recording (capnogram) consisting of multiple individual breath waveforms, each representing a single respiratory cycle of inspiration and expiration. Participants were advised to use the device at home at least two times a day, once in the morning and once in the evening, for a period ranging from six weeks to twelve months (depending on the study). The exact protocol varies by study and is detailed in the supplementary material.

Alongside capnography, concomitant demographic information including sex and age, and clinical data including spirometry, were collected for each participant. Information on specific concomitant data collected for each study can be found in supplementary material.

### Pre-processing and feature engineering

To generate an interpretable machine learning model, each capnogram was translated into a discrete set of features through a similar procedure to that detailed by Talker et al. (2023). These steps are outlined in Fig. 1 and included denoising each capnogram, splitting it into individual breath waveforms, segmenting each breath waveform into phases, and then extracting features from these phases. The capnograph phases are shown in Fig. 2.

**Figure 1.**
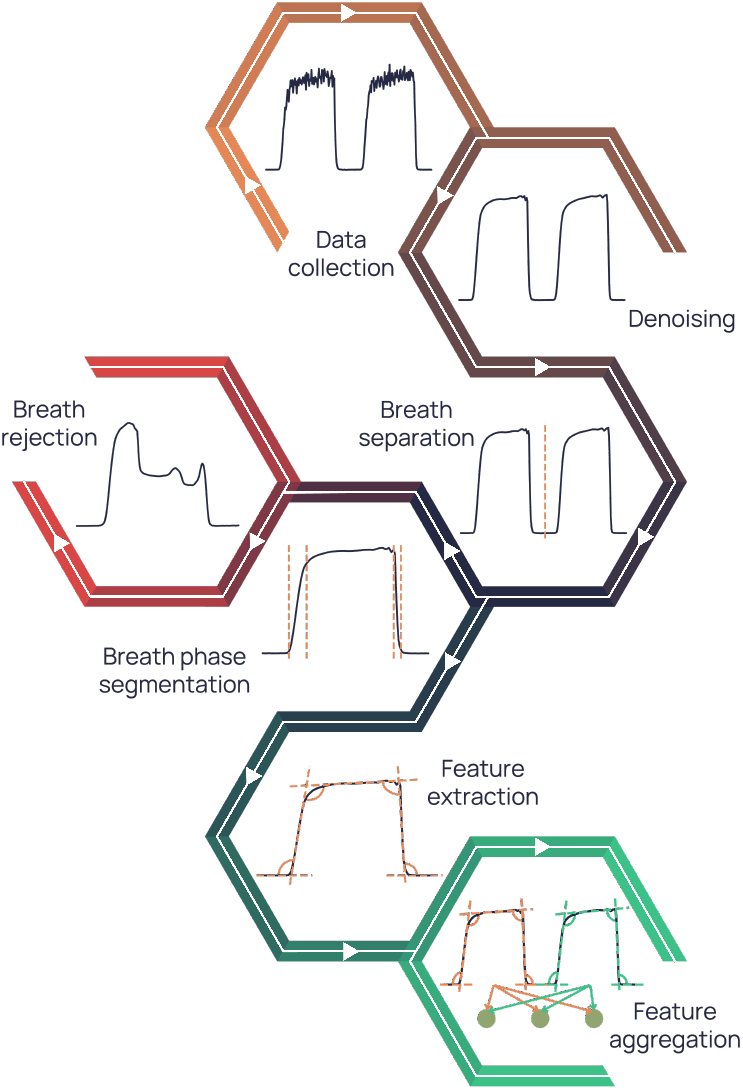
Capnography processing and feature engineering pipeline. Denoising is first applied to collected data followed by separation into individual breaths, before breath phases are segmented and features engineering from these phases. In this pipeline, breaths not meeting the required quality were automatically excluded. Features were then aggregated across all breaths in the final step to get a single set of features for a capnogram.

**Figure 2.**
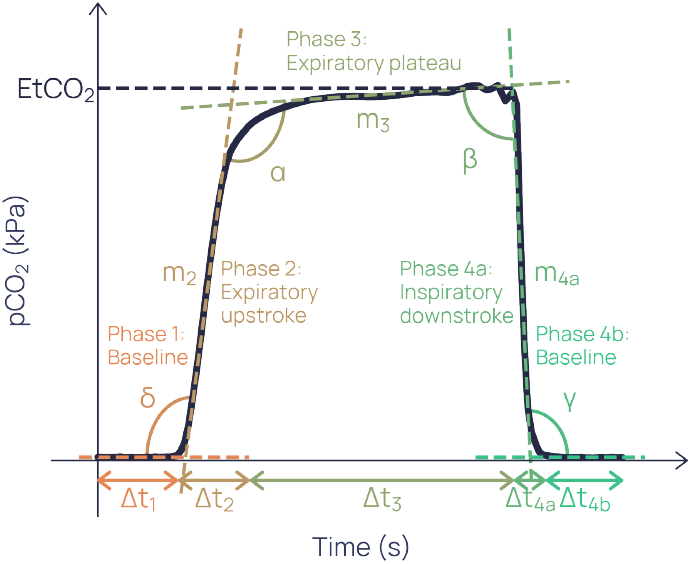
A subset of the breath waveform features. From left-to-right, linear phases are 1, 2, 3, 4a and 4b, with connecting angles *δ, α, β* and *γ*. The duration of the *i*^th^ phase is represented as Δ_*t*_. The transitionary angular regions between linear phases can be considered their own phases. Gradients, *m*_*i*_, can be calculated for the linear phases.

Capnograms that did not meet the required quality criteria (e.g. because of swallowing or nose-breathing artefacts) were excluded. Additionally, to allow the machine learning model to learn the specific pattern of disease associated with asthma, participants with concomitant asthma and at least one cardiorespiratory co-morbidity were excluded. The software and version used for pre-processing and quality control of capnograms was N-Tidal^®^ Diagnose 1 v1.0. N-Tidal^®^ Diagnose 1 v1.0 is a class IIa medical device certified under EU Medical Device Regulations (EU MDR).

In total, 82 features were computed for each capnogram measuring geometry, amplitude, duration, phase gradients and angles between phases (Fig. 2). Many of these features have been previously shown to correspond with airway obstruction in asthma (Hisamuddin et al. 2009; Plantier et al. 2015; You et al. 1994). Each feature was normalised and scaled to a mean of zero and standard deviation of 1 across the dataset.

### Machine learning

The supervised machine learning (ML) methodology implemented for this analysis defined a model that received inputs (each set of 82 features derived from one capnogram) and mathematically transformed them to a prediction (a binary outcome indicating the presence or absence of asthma). The process of model training tuned this prediction to match the participant’s ground truth label (asthma or non-asthma). Large imbalances between the number of samples in each class in the dataset are known to bias ML applications towards low predictive accuracy on the under-represented class (Abd Elrahman et al. 2013). To counter this, a class-balancing procedure was applied that repeatedly removed one capnogram at random from the participant with the most capnograms in the over-represented class (which was asthma). This was repeated until the dataset had an equal number of capnograms in the asthma and non-asthma classes. The machine learning process involved a training stage and a testing stage. The training stage was used to fully optimise a model, which was subsequently evaluated in the testing stage. The training and testing stages required non-overlapping sets of participants to ensure that the ML model was not learning the characteristics of individuals but rather the true underlying signal pertaining to asthma. Thus, before the training stage, the dataset was split into training and testing subsets to ensure any one participant’s records were only in the training stage or the testing stage but not both. Within the training stage, an inner loop was used for hyperparameter optimisation, repeatedly splitting the training set into inner loop training and validation folds using K-fold cross-validation. Approximately 80% of capnograms were kept for the training set, while 20% were reserved for the unseen test set. Furthermore, the dataset was split such that class balance was maintained in both subsets.

A logistic regression (LR) model was trained for the task of asthma diagnostic classification. LR was chosen because it is widely regarded as one of the simplest and most interpretable classical ML models. The interpretability of LR allowed direct visibility of the weights the model was assigning to each input feature. Interpretability in this context is defined as a measure of the transparency of an algorithm’s inner workings, and therefore the ability to understand cause and effect between individual features and the model output. This was imperative to ensure that the model was relying on phases of the waveform that have a plausible physiological basis for being able to identify asthma, without being influenced by possible confounding factors in the training data. Previous work has also shown that linear models such as LR provide similar predictive power to non-linear models when trained on data from the N-Tidal^®^ Handset (Talker et al. 2023).

At the testing stage, the predictions from the unseen test set were compared to their ground-truth labels. Here, model performance metrics were calculated: sensitivity, specificity, positive predictive value (PPV), negative predictive value (NPV), area under the receiver operating characteristic (ROC) curve (AUROC), and average precision from the precision-recall curve. To calculate these, only one capnogram was chosen at random from each participant within the unseen test set to avoid potential bias caused by high accuracy on participants with more records.

### Statistical analysis

The entire ML procedure, from class balancing to testing, was repeated 20 times as part of an outer loop to ensure consistency of findings. The element of randomness in each of these steps meant that the capnograms in the class-balanced dataset and training/test data split were highly unlikely to be the same between iterations. Performance metrics and phase importances provided are the mean of these 20 iterations with confidence intervals as their standard deviation over the 20 iterations. All data analysis and machine learning was performed in Python. Data was handled using the numpy and pandas packages (versions 1.23.5 and 2.0.1 respectively). Models were initialised and trained using the scikit learn package (version 1.2.2).

Demographic bias in the models was assessed by evaluation of diagnostic performance stratified by demographic features that had sufficient diversity - sex and age. Age ranges were chosen to balance the number of participants and capnograms across those ranges. Performance was assessed by individual condition within the non-asthma cohort (COPD, healthy etc.).

Diagnostic instability of physiological testing refers to the variability or inconsistency of test results for the same individual across repeated measurements. It has been well characterised in spirometry (Hei et al. 2020) and is known to be significant for multiple modalities in asthma owing to variable airway obstruction (Lall et al. 2007). To test the longitudinal consistency of ML model outputs, the proportion of participants’ capnograms that were incorrectly predicted were calculated and presented.

### Interpreting capnography feature and phase importances

Importances were attributed to each of the 82 features according to how significantly they contributed to each model’s decisions. For each of the 20 models the absolute values of the weights associated with each feature were ranked, and these ranks averaged over the models. The features were assigned to linear or angular phases of a breath waveform from which they were derived (e.g. phase 1, *α*, etc. as depicted in Fig. 2). The feature importances were aggregated within phases and normalised to yield phase importances.

An average waveform was created for each of the three largest groups (asthma, COPD and healthy) by randomly selecting 5000 capnograms from each group, standardising each of their breaths by time and CO_2_ concentration, and taking the median of all these breaths at each time point.

### Population size justification

To assess whether the study size was sufficient to answer the research question, standard deviation on the mean performance was computed by considering two sources of uncertainty: the variability introduced by the train-test split and the uncertainty arising from the total sample size. Both sources of uncertainty are expected to decrease with more data, but the key question is whether the resulting confidence intervals are narrow enough to answer the research question.

The standard deviation in the model’s performance metrics across the 20 iterations was used to quantify the uncertainty arising from the random, repeated train/test splits. In addition, the standard deviation of a binomial was computed using the size of the unseen test dataset to account for uncertainty due to the finite number of participants in the study. Each source of uncertainty was first combined on the logit scale, where performance metrics are unbounded. The standard deviations were then calculated on this scale and subsequently back-transformed to the probability scale.

## Results

### Baseline characteristics and demographics

80,414 capnograms were collected from 323 study participants and volunteers between 6 December 2015 and 31 January 2022. No adverse events related to the use of the N-Tidal^®^ Handset were reported. Table 1 shows the baseline characteristics and demographic details of participants in the dataset.

**Table 1:**
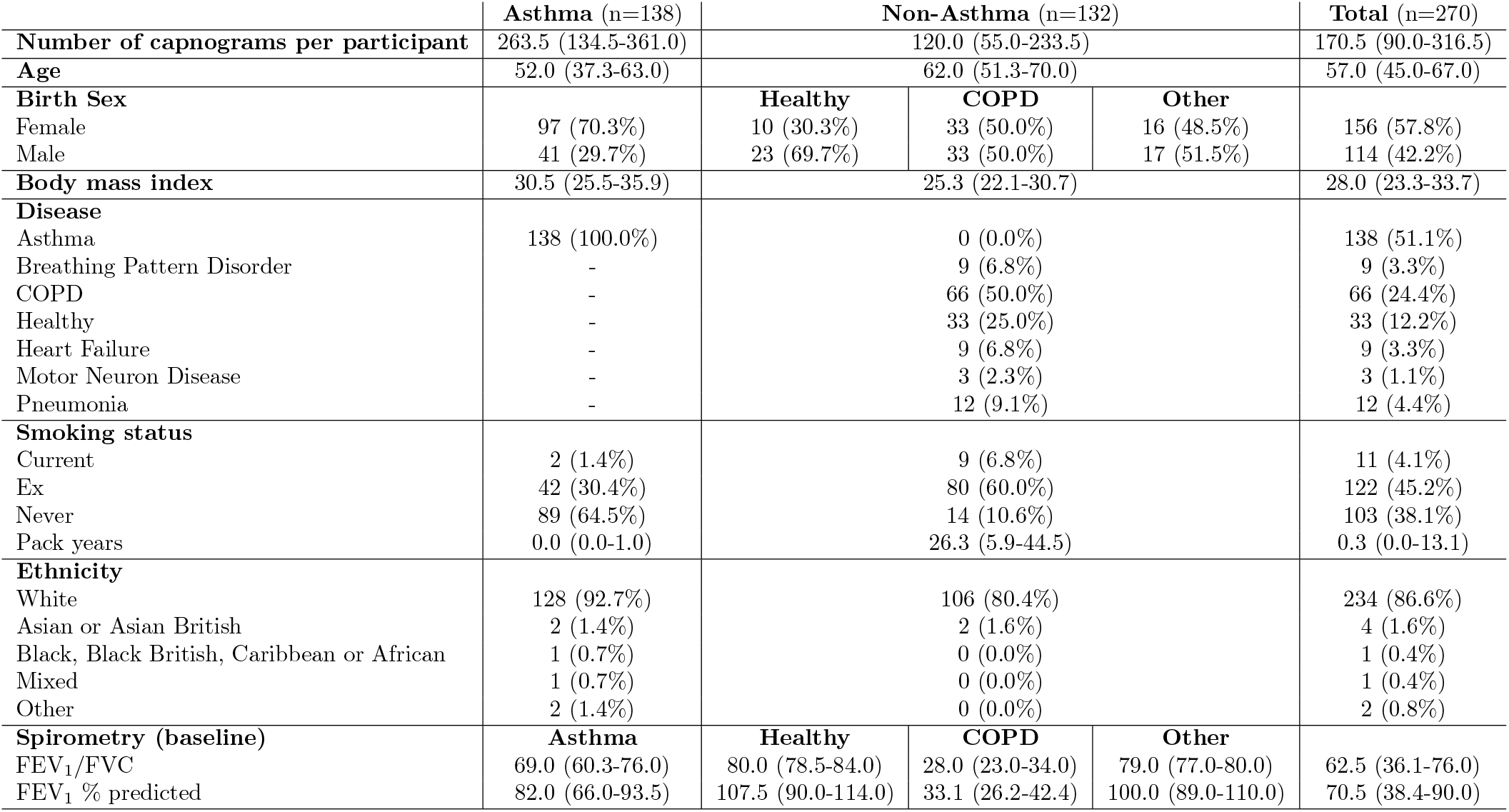
Demographic, medical history, and biomarker breakdown of participants in the dataset used for training and testing. Values are either given as: median (Q1-Q3) or number (percentage of total).

Model pre-processing yielded a quality-controlled dataset with 40,647 asthmatic, and 21,835 nonasthmatic capnograms from 138 individuals with asthma and 132 individuals without asthma respectively. Capnograms that were compromised due to corrupted time stamps, condensation / processing issues, and lacking any processable breaths were discarded to mitigate their impact on the development and evaluation of the classifier.

In the ABRS and GBRS studies, which comprised the asthma cohort, the treatment step of the participant according to the 2019 BTS/SIGN British guidelines (British Thoracic Society 2019) was used as a proxy for disease severity. 4% of asthma participants had no recorded severity, while 5% were at treatment step 1 (regular preventer e.g. low-dose ICS), 22% were at step 2 (initial add-on therapy), 46% were at step 3 (additional controller therapies), and 22% were at step 4 (specialist therapies). The cohort was 70% female, compared with 45% female in the nonasthma cohort. Participants in the asthma cohort were also younger on average, with a mean age of 52 years compared to 62 years in the non-asthma cohort.

COPD was the most frequent alternative diagnosis, accounting for 50% of the non-asthmatic cohort, with the remainder including healthy participants and participants diagnosed with heart failure, pneumonia, or motor neurone disease. Of the participants with a COPD diagnosis, 86% were GOLD 3 or 4, indicating severe or very severe disease. A visualisation of the stages at which participants and capnograms were excluded from the analysis, adapted from the STARD 2015 flow diagram, is shown in Fig. 3, leading to a set of diagnostic outcomes for a randomly selected iteration.

**Figure 3.**
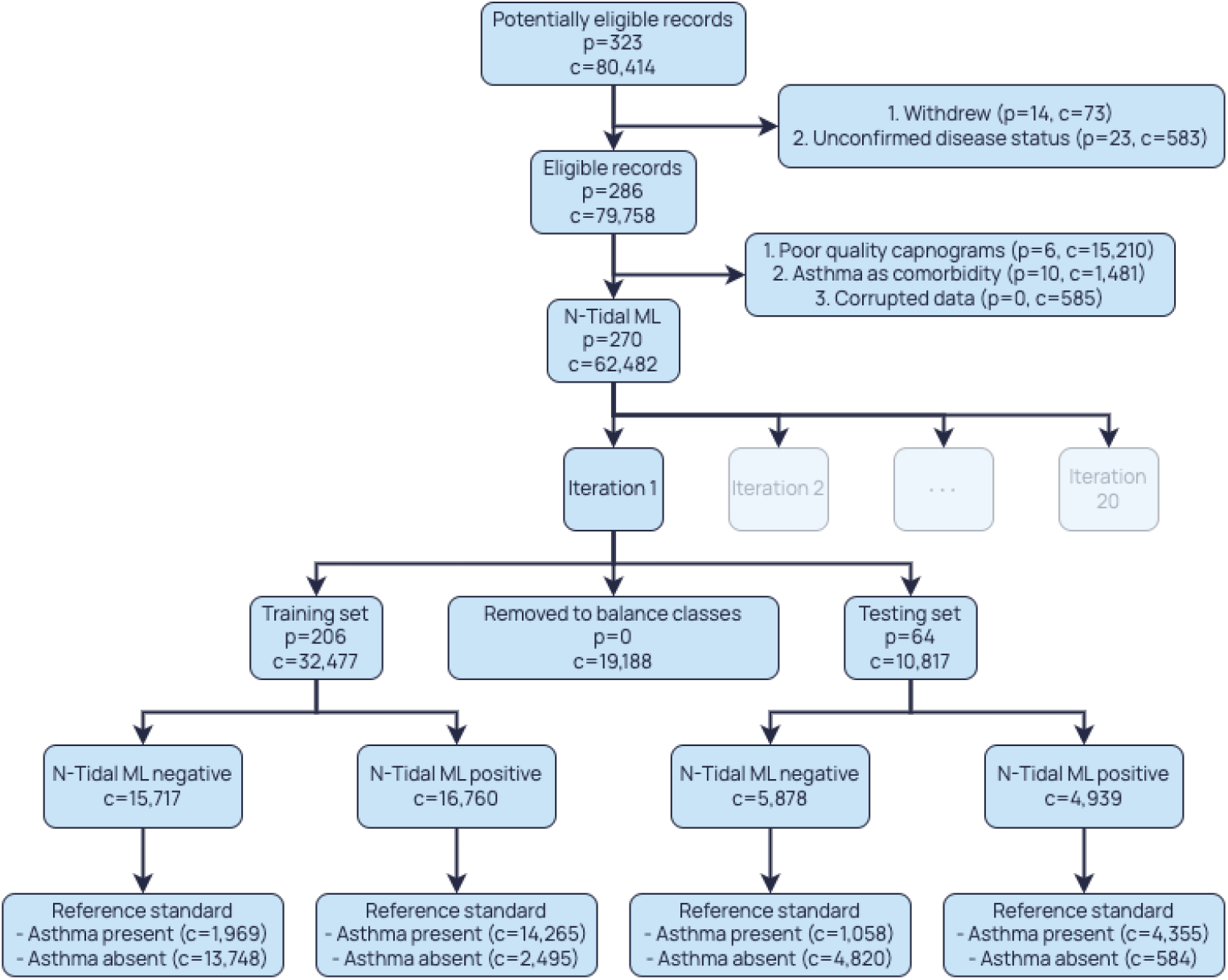
Adapted STARD 2015 flow diagram. Diagnostic test results on the training and test sets are shown for an example iteration. Number of participants = p, number of capnograms = c.

The distribution of the features across the training and unseen test sets can be considered effectively identical. This is because the sets were randomly sampled 20 times, and the performance metrics were aggregated across iterations. With 270 participants, and a 20% test split in each iteration, approximately 98.9% of participants are expected to appear in the unseen test set at least once. Therefore, the distribution of the aggregated unseen test set across all iterations closely approximates the distribution of the full dataset, and by extension, that of the training set.

### Machine learning classification performance

On the unseen test set, the logistic regression models achieved an AUROC of 0.910 ± 0.027 and a precision of 90.5 ± 3.4% (Fig. 4). The models also demonstrated a mean sensitivity of 83.3 ± 3.8%, specificity of 85.2 ± 5.6%, PPV of 87.4 ± 4.3%, and NPV of 81.0 ± 3.6% (Fig. 5) in diagnosing asthma.

**Figure 4.**
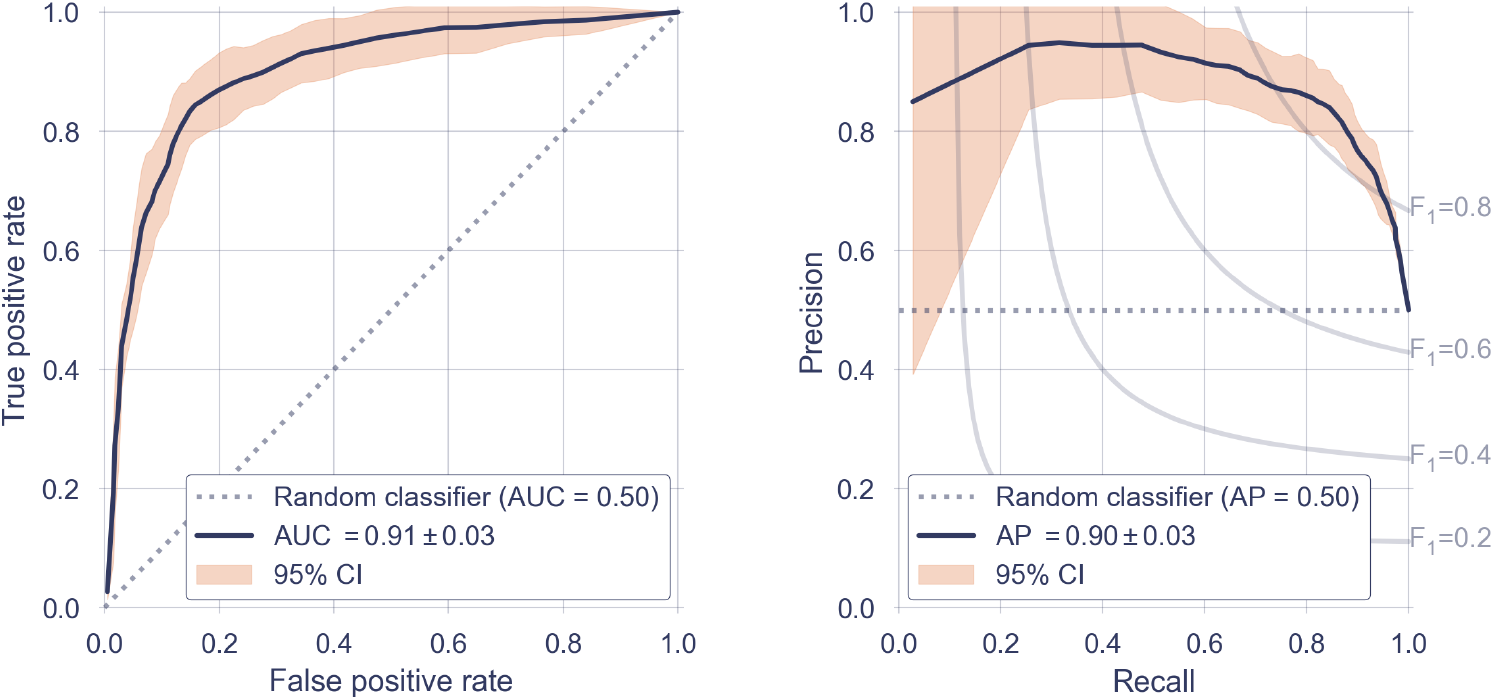
(a) Receiver operator characteristic (ROC) curve, reported with results of a theoretical ‘random’ classifier (dotted line) with no predictive power, and a 95% confidence interval, calculated across all 20 iterations. (b) Precision-Recall Curve, reported with the results of a theoretical ‘random classifier’ (dotted line) and the average precision (AP).

**Figure 5.**
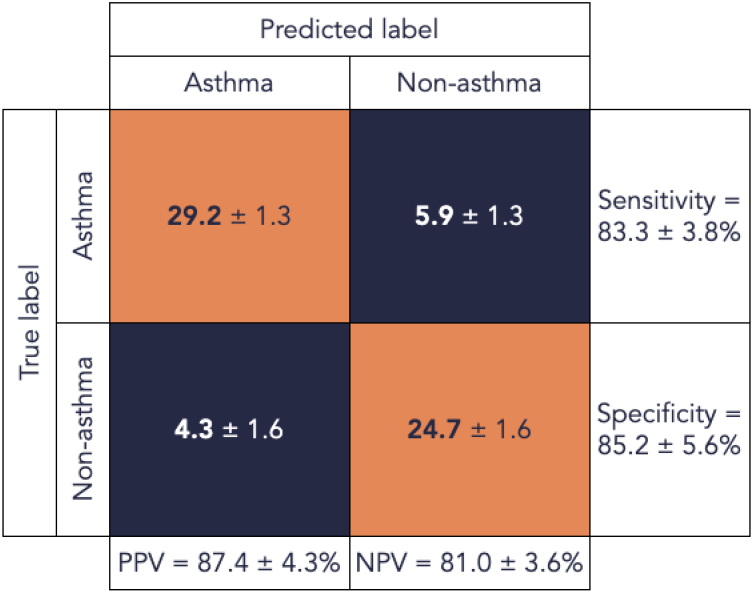
Confusion matrix showing classification performance (predicted label) against the clinical diagnosis (true label). Performance is reported in terms of the average number of participants in the test set that fell in each quadrant.

### Phase Importances

To illustrate the phases of the capnogram that were determined to be most important in driving asthma diagnosis by the logistic regression model, the phase importances were calculated from feature importances (Fig. 6). The transitional alpha and beta angles are highlighted as the most discriminative regions, with importance decreasing towards the baselines.

**Figure 6.**
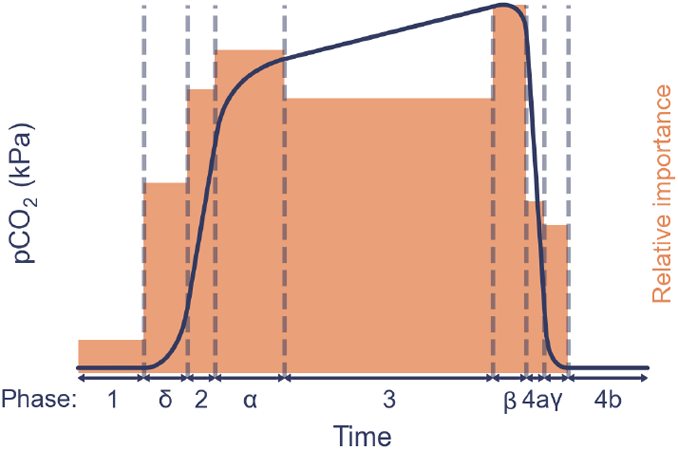
Relative importance for each phase of the waveform in the models’ decision-making, imposed on an example average waveform.

Next, the average waveform was calculated for each group (Fig. 7). Individuals with asthma exhibited a more gradual incline of the expiratory upstroke and a steeper expiratory plateau compared to the healthy waveform, and showed less pronounced curvature of the alpha angle region compared to individuals with COPD.

**Figure 7.**
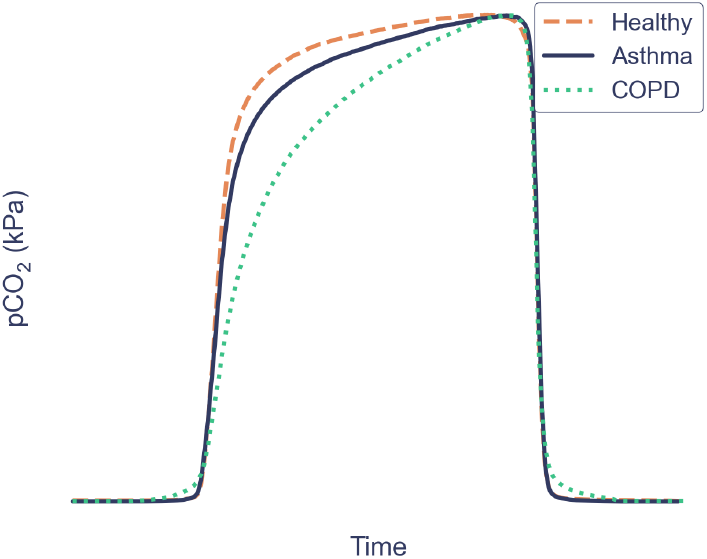
Average waveforms calculated for the COPD, asthma and healthy groups.

### Model bias

To evaluate whether any model bias was present, the mean accuracy (± standard deviation over 20 iterations) within demographic and disease categories was calculated on the test set. Note that the number of capnograms for each cohort in the unseen test set varied between iterations, hence the standard deviation for this column.

The difference in accuracy between sexes was minimal and within 1 standard deviation, however an increase in prediction accuracy was observed as age increased. The highest accuracy for non-asthma prediction was observed in individuals with COPD; reduced mean accuracy and greater variability across iterations was noted amongst the healthy and asthma cohorts. All demographic groups demonstrated an accuracy exceeding 80% (see Table 2).

**Table 2:**
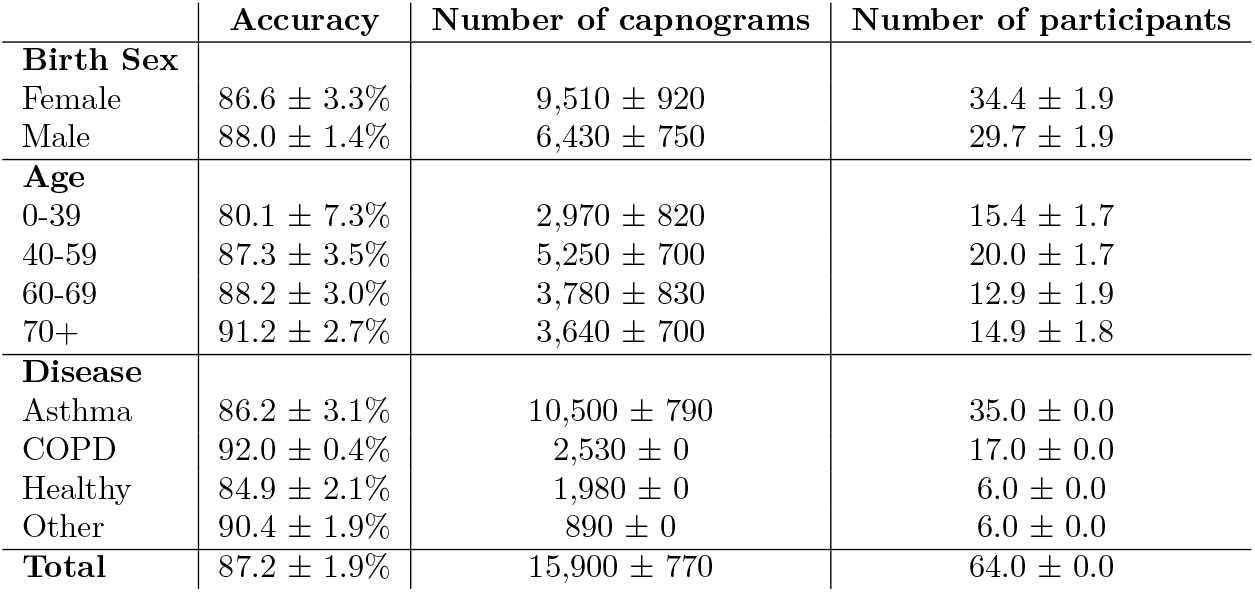
Classification accuracy stratified by sex, age and disease group. Values are given as mean ± standard deviation over 20 iterations.

### Prediction consistency

Next, misclassified waveforms were interrogated to determine the geometric characteristics of waveforms that were both correctly and incorrectly classified. Average breath waveforms from individual asthma, healthy and COPD capnograms, predicted with varying probabilities of being asthmatic, are shown in Fig. 8. The prediction probabilities were calculated as the mean probability of asthma according to each of the 20 iterations, using only the capnograms in the unseen test the unseen test set for that specific iteration. Participants with an underlying clinical diagnosis of COPD who were predicted by the LR models as having asthma had waveform geometries more similar to the asthma group. Participants with a clinical diagnosis of asthma who were predicted by the LR models as not having asthma had relatively flat expiratory plateau phases (similar to healthy participants). Participants who were healthy, but who were predicted by the LR models as having asthma, had an incline on the expiratory plateau phase.

**Figure 8.**
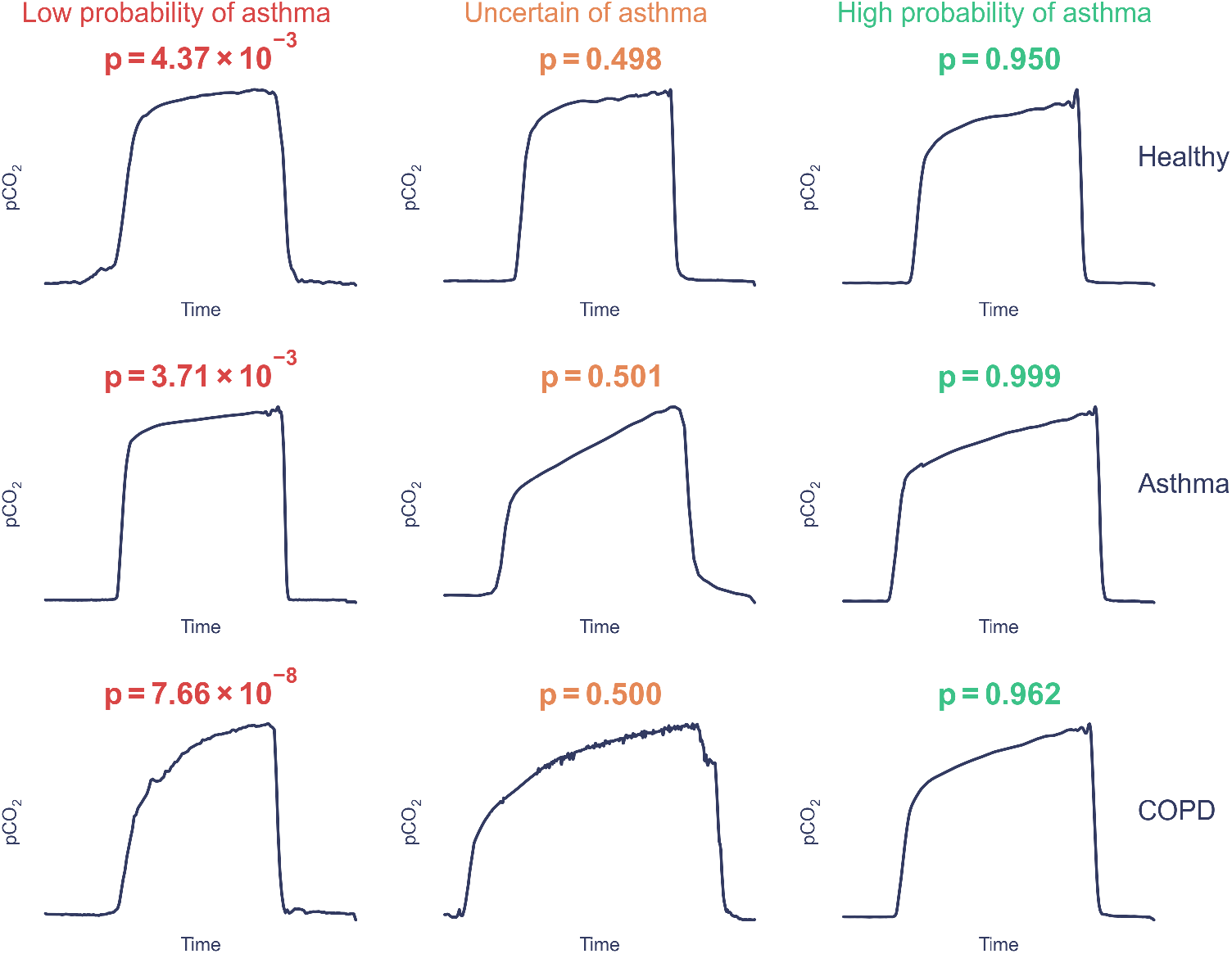
Average breath waveforms of capnograms with varying prediction probabilities of asthma. Each row shows examples from participants with different primary clinical diagnoses. The leftmost column shows examples of waveforms that the LR models determined were less likely to be asthma, the rightmost column shows examples of waveforms that the LR models determined most likely to be asthma, and the central column shows uncertain predictions. The average probability of having asthma across the 20 iterations is denoted by p.

Finally, analysis of the consistency of ML model predictions (by evaluating model performance on repeat measurements for individual participants in each unseen test set test set) showed that the median proportion of incorrect predictions per participant was 4.2% (Fig. 9). Only 7.5% of participants were incorrectly classified more than 50% of the time, demonstrating high consistency of prediction over multiple repeat tests and even over prolonged periods of time.

**Figure 9.**
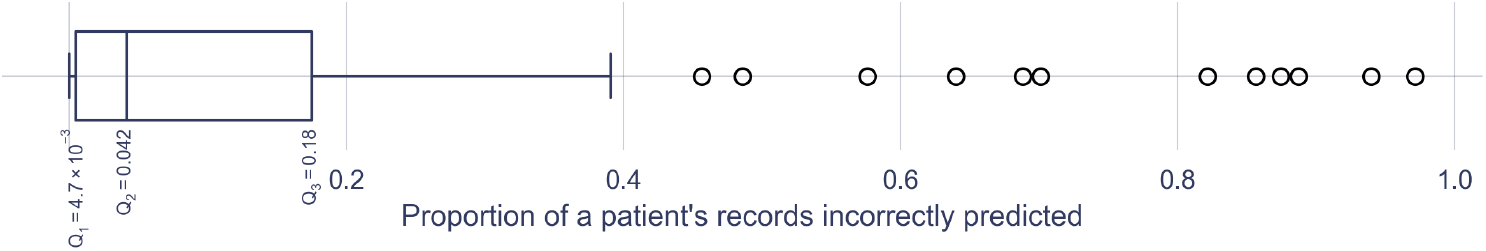
Proportion of each test set participants’ capnograms that were incorrectly classified.

### Population Size Justification

While the unseen test set dataset in a single iteration was relatively small, resulting in a large standard deviation in the performance metrics, the repeated random sampling across 20 iterations effectively increased the number of participants contributing to the evaluation to 98.9% of the total dataset. The effective test dataset included 136 participants with asthma and 130 participants without asthma. Accounting for both variability across iterations and finite-sample uncertainty (unlike the performance statistics presented in Machine Learning Classification performance, which present variability across iterations within the current dataset), the model sensitivity was 83.3% ± 5.1%, and the specificity was 85.2% ± 6.7%. The approach used was conservative as it assumed independence in the sources of uncertainty, and therefore these results demonstrate the strong potential of the presented model in diagnostic classification of asthma.

## Discussion

In this manuscript, we describe the development and diagnostic performance of a potential novel diagnostic classifier for asthma using the N-Tidal^®^ handheld capnometer (N-Tidal^®^ Handset) combined with a logistic regression classifier, in participants with and without asthma. The classifier demonstrated high diagnostic PPV (87.4 ± 4.3%) and NPV (83.3 ± 3.8%). The analysis of prediction variability in individual participants revealed that the median number of incorrectly classified capnograms per participant was 4.2%, indicating a high level of consistency across repeat readings and over time, with most participants receiving a correct diagnosis in over 95% of successful breath recordings.

The waveforms in Fig. 7 illustrate that participants with asthma exhibit a more gradual incline of the expiratory upstroke and a steeper expiratory plateau compared to the healthy waveform. While a longer and more curved alpha angle is seen to some extent in asthma, it is most clearly present in the average COPD waveform. Healthy waveforms do not display these alpha angle changes.

The process of waveform averaging to generate the comparisons in Fig. 7 does suppress components of individual breaths that may vary in time or CO_2_ concentration. Therefore, the models may have picked up on features that are not apparent from this visual display alone. To attempt to highlight some of this ambiguity, Fig. 8 visualises average breath waveforms from individual capnograms. The second row of capnograms from asthmatic participants, alongside the third column of capnograms that were predicted as most likely asthmatic, highlights that the significant drivers of an asthma diagnosis (according to the model) were a greater alpha angle and expiratory plateau tangent. The analysis supports the physiological plausibility that these features characterise the inhomogeneity of airflow obstruction and ventilation-perfusion ratios across different lung regions, resulting from the narrowing of distal airways due to structural and inflammatory changes (Almeida et al. 2011). Feature importances are displayed in Fig. 6 and corroborate the importance of the alpha and expiratory plateau regions. The beta angle region (both the beta angle itself and the magnitude of pCO_2_) was also a significant driver of classification.

Importantly, the training and unseen test sets include participants who were established on treatment for their respective conditions. The significant majority of participants with asthma were prescribed long-acting bronchodilators and inhaled corticosteroids while those with COPD were predominantly prescribed at least one longacting bronchodilator. The diagnostic performance in the context of inhaled therapy prescription is advantageous given participants attending for diagnostic testing may not be treatment naïve despite instructions, creating diagnostic uncertainty using existing modalities like spirometry and FeNO (NICE 2024b). The waveforms in Fig. 7 show that even in a treated patient population, there are changes in the CO_2_ waveform of participants with asthma that are likely to reflect features of chronic inflammation and remnant inhomogeneous airflow obstruction.

Additionally, this research highlights the possibility of quantifying airway changes associated with asthma from a single breath recording, potentially reducing the need for reversibility testing. This has significant implications for clinical practice where the availability of a skilled, trained workforce to carry out reversibility testing with spirometry is limited; the results in this manuscript demonstrate the feasibility of developing a point-of-care diagnostic test with a sensitivity that far surpasses the sensitivity of first-line diagnostic tests within the UK asthma pathway.

The waveforms in Fig. 8 highlight that misclassifications occurred where waveform geometry was more similar to that of a different clinical diagnosis. For example, participants with a clinical diagnosis of COPD who were misclassified as having asthma had waveform geometries more consistent with asthma: a relatively preserved uncurved alpha angle transition phase with an incline on the expiratory plateau. Equally, participants with asthma misclassified as not having asthma had waveform geometries that were less similar to the asthma group, e.g., with relatively flat expiratory plateau which would be more consistent with a healthy individual. While some participants in the dataset may have been labelled incorrectly because of the limitations of current diagnostic testing, it is also possible that more work is needed to understand and characterise the diverse waveform shapes observed in asthma and healthy participants in more detail.

It is well established that asthma exhibits sexual dimorphism, with higher prevalence and severity in adult females than in adult males (Reddy et al. 2023). Differences in the physiological signal measured by the capnograph may therefore be expected. However, predictive performance between sexes demonstrated similar accuracies between males and females. Stratifying performance by age category showed improved performance with increasing age. The youngest age category of 0-39 yielded the lowest mean accuracy of 80.1 ± 7.3%. This may be due to having fewer participants within this group, containing only ~ 5 participants per decade, compared to ~ 13 participants per decade in the 60-69 group. The prevalence of COPD was also higher in the older age groups, therefore inflating model accuracy because the diagnostic performance of the model was notably higher for COPD compared with asthma. Furthermore, disease was more severe within the older age groups, most likely making classification easier. Mean FEV_1_ % predicted for participants younger than 40 was 93%, while it was 54% for participants older than 60. This observation aligns with findings from Zein et al. (2015), which showed that, among participants with asthma, the probability of severe asthma increased with age, and that those over 45 were 2.73 times more likely to have severe asthma than those younger than 45.

Since most non-asthma participants in this paper had a primary diagnosis of COPD there was the possibility that the classifier had primarily learnt the signal separating asthma and COPD, instead of the signal that could differentiate asthma from a variety of differential diagnoses. However, the predictive accuracy of 84.9 ± 2.1% amongst the healthy participants and 90.4 ± 1.9% amongst other non-asthma conditions indicates that the models were able to effectively identify conditions other than asthma and COPD as non-asthma. To support this conclusion, a greater sample size of non-COPD non-asthma participants would be beneficial.

### Limitations and future work

The asthma and COPD participants within this study mainly consisted of those on the severe ends of their respective spectrums. It is possible that it would be more challenging to distinguish milder COPD and asthma, and to distinguish milder asthma from healthy. Future research should validate these findings in a cohort containing participants with milder disease. 87% of the dataset came from a white ethnic background, and therefore there was not enough data in this dataset to conclusively validate performance in non-white groups. Including more healthy participants and those with other cardiorespiratory conditions that may present similarly to asthma would improve confidence in model specificity. Furthermore, the age of the asthma group was quite high, consisting predominantly of adults, with only two participants under the age of 18, which may limit the generalisability to younger individuals with suspected asthma. Further data collection is required to evaluate the diagnostic model performance in a cohort of more diverse ages, ethnicities, and severities of disease. This work also used data from participants with prior diagnoses of asthma and non-asthmatic conditions, who had been prescribed appropriate therapy. While also a strength of this work, future work should investigate performance in a cohort with suspected asthma (i.e. an undifferentiated population, such as one that might be seen in a primary care diagnostic setting). In addition, the dataset used in this study was quality-controlled to eliminate the impact of erroneous capnograms on the development and evaluation of the model. Future work should therefore investigate the usability of the model in clinical settings to determine how many capnogram recordings result in an analysable output.

To further improve model performance, there may be benefit in evaluating changes in capnography features over time, given that asthma is known to cause variable airway obstruction (Lall et al. 2007). There may also be benefit from use of higher complexity ML models.

### Conclusions

This research presents an accurate and highly interpretable diagnostic model able to classify asthma using a single breath recording captured with the N-Tidal^®^ capnometer (N-Tidal^®^ Handset). Applied to new breath recordings, this algorithm has the potential to provide point-of-care diagnosis for asthma. The diagnostic classifier presented here achieved high diagnostic precision and recall, with the majority of participants receiving the same diagnosis in over 95% of their successful breath recordings. There was low variability in classification accuracy between iterations and demographic groups, but further data is required in those with milder asthma and COPD, those of non-white ethnicity, and those with a greater diversity of cardiorespiratory conditions. In contrast to commonly used ‘black box’ machine learning methodologies, a set of highly explainable methods were used that could provide traceability back to individual features of the CO_2_ waveform, and the associated physiological properties that are suggestive of obstructive airways disease.

The portability and ease of use of capnography could make its use, in combination with the type of diagnostic algorithms generated in this paper, an attractive community diagnostic test for patients and healthcare systems. Further model development and assessment of diagnostic accuracy is required to ensure that it fulfils its potential to benefit clinical practice and improve patient care.

## Data Availability

The datasets generated during and/or analysed, and the algorithms developed, during the current study are not publicly available for data protection, confidentiality, and commercial sensitivity reasons.

## Declarations

### Ethics approval and consent to participate

Ethical approval was obtained from the South Central - Berkshire Research Ethics Committee (REC) for GBRS and ABRS, the Yorkshire and the Humber REC for CBRS and the West Midlands Solihull REC for CBRS2. All participants provided written informed consent to participate; further details can be found in the clinical trials section.

### Public Involvement

There was no public or patient involvement in the design, conduct, reporting, interpretation, or dissemination of the study.

## References

Aaron, Shawn D. et al. (2018). “Underdiagnosis and Overdiagnosis of Asthma.” In: American Journal of Respiratory and Critical Care Medicine 198.8, pp. 1012–1020. ISSN: 1535-4970. doi: 10.1164/RCCM.201804-0682CI. url: https://pubmed.ncbi.nlm.nih.gov/29756989/.

Abd Elrahman, Shaza M. et al. (2013). “A Review of Class Imbalance Problem.” In: Journal of Network and Innovative Computing 1, pp. 332–340.

Almeida, Celize C. B. et al. (2011). “Volumetric capnography to detect ventilation inhomogeneity in children and adolescents with controlled persistent asthma.” In: Jornal de Pediatria 87.2, pp. 163–168. ISSN: 1678-4782. doi: 10.2223/JPED.2077. url: https://pubmed.ncbi.nlm.nih.gov/21503374/.

British Thoracic Society (2019). SIGN 158 British Guideline on the Management of Asthma: Quick Reference Guide.url: https://www.brit-thoracic.org.uk/document-library/guidelines/asthma/btssign-asthma-guideline-quick-reference-guide-2019/.

Cohen, Jeremie F. et al. (2016). “STARD 2015 guidelines for reporting diagnostic accuracy studies: explanation and elaboration.” In: BMJ Open 6.11. ISSN: 2044-6055. doi: 10.1136/bmjopen-2016-012799. url: https://bmjopen.bmj.com/content/6/11/e012799.

Collins, G. S. et al. (2024). “TRIPOD+AI statement: updated guidance for reporting clinical prediction models that use regression or machine learning methods.” In: BMJ 385, e078378. doi: 10.1136/bmj-2023-078378.

Hei, S. J. van de et al. (2020). “Quality of spirometry and related diagnosis in primary care with a focus on clinical use.” In: npj Primary Care Respiratory Medicine 30.1, pp. 1–7. ISSN: 2055-1010. doi: 10.1038/s41533-020-0177-z. url: https://www.nature.com/articles/s41533-020-0177-z.

Hisamuddin, N. A. R. Nik et al. (2009). “Correlations between capnographic waveforms and peak flow meter measurement in emergency department management of asthma.” In: International Journal of Emergency Medicine 2.2, p. 83. ISSN: 1865-1372. doi: 10.1007/s12245-009-0088-9. url: https://www.ncbi.nlm.nih.gov/pmc/articles/PMC2700227/.

Howard, S. (2023). ““Silent scandal” of missing lung diagnostics in England’s most deprived areas—where respiratory disease is most prevalent.” In: BMJ 382, p. 2140. doi: 10.1136/bmj.p2140.

Jaffe, Michael B. (2017). “Using the features of the time and volumetric capnogram for classification and prediction.” In: Journal of Clinical Monitoring and Computing 31.1, pp. 19–41. ISSN: 1573-2614. doi: 10.1007/s10877-016-9830-z. url: https://pubmed.ncbi.nlm.nih.gov/26780902/.

Kupczyk, M. et al. (2010). “Reduction of asthma burden is possible through National Asthma Plans.” In: Allergy 65.4, pp. 415–419. ISSN: 1398-9995. doi: 10.1111/j.1398-9995.2009.02265.x. url:https://pubmed.ncbi.nlm.nih.gov/20102359/

Lall, C. A. et al. (2007). “Airway resistance variability and response to bronchodilator in children with asthma.” In: European Respiratory Journal 30.2, pp. 260–268. ISSN: 0903-1936. doi: 10.1183/09031936.00064006.

Mieloszyk, R. J. et al. (2014). “Automated Quantitative Analysis of Capnogram Shape for COPD–Normal and COPD–CHF Classification.” In: IEEE Transactions on Biomedical Engineering 61.12, pp. 2882–2890. doi: 10.1109/TBME.2014.2332954.

Mou, Xiuying et al. (2024). “A Novel Approach for the Detection and Severity Grading of Chronic Obstructive Pulmonary Disease Based on Transformed Volumetric Capnography.” In: Bioengineering 11.6, p. 530. doi: 10.3390/bioengineering11060530. url: https://doi.org/10.3390/bioengineering11060530.

Mukherjee, Mome et al. (2016). “The epidemiology, healthcare and societal burden and costs of asthma in the UK and its member nations: Analyses of standalone and linked national databases.” In: BMC Medicine 14.1, pp. 1–15. ISSN: 1741-7015. doi: 10.1186/s12916-016-0657-8. url: https://bmcmedicine.biomedcentral.com/articles/10.1186/s12916-016-0657-8.

NHS (2021). Asthma - NHS. url: https://www.nhs.uk/conditions/asthma/.

NHS (2024). Commissioning standards for spirometry. url: https://www.england.nhs.uk/long-read/commissioning-standards-for-spirometry/.

NICE (2023). When should I suspect asthma? url: https://cks.nice.org.uk/topics/asthma/diagnosis/diagnosis/.

NICE(2024a). Asthma pathway (BTS, NICE, SIGN). url: https://www.nice.org.uk/guidance/ng244.

NICE(2024b). Asthma: diagnosis, monitoring and chronic asthma management (update). url: https://www.nice.org.uk/guidance/ng245/resources/asthma-diagnosis-monitoring-and-chronic-asthma-management-bts-nice-sign-pdf-66143958279109.

NICE (2024c). Cost-utility analysis: Most costeffective sequence or combination of tests to diagnose asthma. url: https://www.nice.org.uk/guidance/ng245/evidence/cost-utility-analysis-most-costeffective-sequence-or-combination-of-tests-to-diagnose-asthma-pdf-13558289293/.

NICE (2024d). Evidence reviews for bronchial challenge with histamine and methacholine for the diagnosis of asthma. url: https://www.nice.org.uk/guidance/ng245/evidence/h-bronchial-challenge-with-histamine-and-methacholine-for-the-diagnosis-of-asthma-pdf-13558146741.

Pavord, I. D. et al. (2018). “After asthma: redefining airways diseases.” In: Lancet 391.10118, pp. 350– 400. doi: 10.1016/S0140-6736(17)30879-6. url: https://pubmed.ncbi.nlm.nih.gov/28911920/.

Pertzov, B. et al. (2021). “Use of capnography for prediction of obstruction severity in non-intubated COPD and asthma patients.” In: Respiratory Research 22.1, p. 154. doi: 10.1186/s12931-021-01747-3. url: https://www.ncbi.nlm.nih.gov/pmc/articles/PMC8138110/.

Plantier, Laurent et al. (2015). “Methacholine-Induced Variations in Airway Volume and the Slope of the Alveolar Capnogram Are Distinctly Associated with Airflow Limitation and Airway Closure.” In: PLoS ONE 10.11, e0143550. ISSN: 1932-6203. doi: 10.1371/journal.pone.0143550. url: https://www.ncbi.nlm.nih.gov/pmc/articles/PMC4658077/.

Reddy, K. D. et al. (2023). “Sexual dimorphism in chronic respiratory diseases.” In: Cell Biosci 13.1, p. 47. doi: 10.1186/s13578-023-00998-5. url: https://www.ncbi.nlm.nih.gov/pmc/articles/PMC10029954/.

Simpson, Andrew J. et al. (2024). “Asthma diagnosis: a comparison of established diagnostic guidelines in adults with respiratory symptoms.” In: Lancet eClinicalMedicine 76, p. 102813. doi: 10.1016/j.eclinm.2024.102813.

Singh, Om Prakash et al. (2018). “Automatic Quantitative Analysis of Human Respired Carbon Dioxide Waveform for Asthma and Non-Asthma Classification Using Support Vector Machine.” In: IEEE Access 6, pp. 55245–55256. ISSN: 2169-3536. doi: 10.1109/ACCESS.2018.2871091.

Sinyor, Benjamin et al. (2023). Pathophysiology of Asthma. url: https://www.ncbi.nlm.nih.gov/books/NBK551579/.

Talker, Leeran et al. (2023). “Machine diagnosis of chronic obstructive pulmonary disease using a novel fast-response capnometer.” In: Respiratory Research 24.1, pp. 1–11. ISSN: 1465-993X. doi: 10.1186/s12931-023-02460-z. url: https://respiratory-research.biomedcentral.com/articles/10.1186/s12931-023-02460-z.

WHO (2023). Asthma. url: https://www.who.int/news-room/fact-sheets/detail/asthma.

You, B. et al. (1994). “Expiratory capnography in asthma: evaluation of various shape indices.” In: European Respiratory Journal 7.2, pp. 318–323. ISSN: 0903-1936.

Zein, Joe G. et al. (2015). “Asthma Is More Severe in Older Adults.” In: PLoS ONE 10.7, e0133490. ISSN: 1932-6203. doi: 10.1371/journal.pone.0133490. url: https://www.ncbi.nlm.nih.gov/pmc/articles/PMC4511639/.

