## Supplementary material for "Development and evaluation of a multivariate prediction model for diagnosing asthma in patients with clinically suspected asthma using capnography"

Henry Broomfield[1], Leeran Talker[1], Rui Hen Lim[1], Joseph Massingham[1], Gabriel Lambert[1], Helen Ashdown[2,3], Gail Hayward[2,3], Daniel Neville[4], Thomas Brown[4,5], Laura Wiffen[4,5], Anoop Chauhan[4,5], and Ameera Patel[1]

[1] *TidalSense Limited, Cambridge, UK*
[2] *Nuffield Department of Primary Care Health Sciences, University of Oxford, Oxford, UK*
[3] *NIHR HealthTech Research Centre in Community Healthcare, Oxford Health NHS Foundation Trust, Oxford, UK*
[4] *Portsmouth Hospitals University NHS Foundation Trust, Portsmouth, UK*
[5] *Faculty of Science and Health, University of Portsmouth, Portsmouth, UK*

### Clinical Studies

| **Study** | **COPD Breathing Record Study (CBRS)** | **General Breathing Record Study (GBRS)** | **COPD Breathing Record Study 2 (CBRS2)** | **Asthma Breathing Record Study (ABRS)** |
|---|---|---|---|---|
| **Primary Objective** | To collect a longitudinal observation study database of capnograph records for up to 30 participants with recent or recurrent exacerbations of COPD over 6 weeks. | To explore the characteristics of the $TBCO_2$ waveform that can differentiate between different respiratory and cardiac conditions and establish a profile for healthy controls. | To assess the changes in key parameters in the $TBCO_2$ waveform, including the $\alpha$ angle, the minimum $CO_2$ level achieved and the stability of the expiratory cycle, during the transition from stable COPD to during acute exacerbations. | To determine characteristics within the $TBCO_2$ waveform shape that identify deteriorations in respiratory condition and discriminate between poorly and well-controlled asthma. |

Continued on next page

Table 1 – continued from previous page

| Study | CBRS | GBRS | CBRS2 | ABRS |
|---|---|---|---|---|
| **Population Inclusion Criteria** | Males or females >18 years old with COPD diagnosis from two groups: community group (chronically elevated $PaCO_2$, frequent exacerbations) and acute admissions group (hospitalised with a COPD exacerbation). | Disease cohorts: males or females ≥16 years old selected from hospital outpatient clinic lists or inpatient wards. Includes healthy volunteers. Asthma: clinician diagnosis >6 months with evidence of variability, reversibility or hyper-responsiveness; BTS Stage 3–5; ≥2 exacerbations in 12 months; ≥1 exacerbation in 6 months; exacerbation-free for 2 weeks. Healthy: no diagnosis or treatment for lung, cardiac or neuromuscular conditions; BMI ≤40; smokers ≤5 pack-years. Other: confirmed clinical diagnosis. | Males or females >40 years old with a diagnosis of COPD and at least one moderate exacerbation in the previous year. | Males or females ≥7 years old with clinician-confirmed moderate/severe asthma (BTS 2–5), ACQ>1, and >1 exacerbation in <12 months. |
| **Population Exclusion Criteria** | Neuromuscular disorders or kyphoscoliosis. | Comorbidities affecting spirometry or other lung function measurements. | Neuromuscular disorders or kyphoscoliosis; exacerbation requiring antibiotics or corticosteroids <2 weeks before start; comorbidities affecting lung function. | Comorbidities affecting lung function (including BPD, COPD); smokers (current or ex) >10 pack-years. |
| **Clinical condition(s)** | COPD | Asthma, breathing pattern disorder, chronic heart failure, motor neuron disease, pneumonia, healthy volunteers | COPD | Asthma |
| **Number of participants** | COPD: 30 | Asthma: 20; BPD/ CHF/ MND/ Pneumonia/ Healthy: 10 each; Total: 70 | COPD: 50 | Asthma: 124 (92 from primary care, 32 from secondary care). |

Continued on next page

Table 1 – continued from previous page

| **Study** | **CBRS** | **GBRS** | **CBRS2** | **ABRS** |
|---|---|---|---|---|
| **Location** | Addenbrookes Hospital, Cambridge | Queen Alexandra Hospital and specialist secondary-care community clinics, Portsmouth | COPD Centre, Addenbrookes Hospital, Cambridge | Queen Alexandra Hospital, Portsmouth and GP practices, Oxford |
| **Recruitment setting** | Outpatient, inpatient | Outpatient | Outpatient | Outpatient, inpatient, primary care |
| **Duration** | 17 Feb 2016 – Dec 2016 | 9 Aug 2017 – 4 Jul 2018 | 15 Aug 2017 – 23 Nov 2018 | 11 Feb 2020 – 31 Jan 2022 |
| **Clinical Trials.gov Identifier** | NCT02814253 | NCT03356288 | NCT03615365 | NCT04504838 |
| **Number of capnograms** | 2620 | 15803 | 14885 | 38026 |
| **Additional data collected** | Medical history, clinical assessment, demographics, vital signs, spirometry, routine blood tests, blood gases. | Medical history, clinical assessment, demographics, vital signs, spirometry (not HF or pneumonia groups), and disease-specific assessments. | Medical history, clinical assessment, demographics, symptoms, vital signs, spirometry. | Medical history, clinical assessment, demographics, vital signs, spirometry (plus FeNO, oscillometry, full body plethysmography), routine blood tests. |
| **Device use relative to inhaler use** | Not specified | Not specified | Not specified | Device used before inhaler use |
| **Treatments given** | Each patient prescribed at least one long-acting bronchodilator. | Asthma: majority prescribed corticosteroids + long-acting bronchodilators. Other conditions: condition-specific. | Each patient prescribed at least one long-acting bronchodilator. | Majority prescribed corticosteroids + long-acting bronchodilators. |

Table 1: Summary of the four clinical studies from which the paper has drawn its data.

### Schedules of Assessment

Table 2: Schedule of assessments for COPD Breathing Record Study (CBRS).

| | **Screening** | **Monitoring Period** | | | | **Follow-up** |
|---|---|---|---|---|---|---|
| | | **Week 0** | **Week 2** | **Week 4** | **Week 6** | **Week 8** |
| Window (days) | -14 to 0 | | ±4 | ±4 | ±3 | ±7 |
| **Study visit** | **1**[1] | **2**[1] | **3** | **4** | **5** | **6** |
| *Location* | *Home, clinic or hospital* | *Home, clinic or hospital* | *Home* | *Home* | *Clinic* | *Telephone* |
| Informed consent | X | | | | | |
| Demography | X | | | | | |
| Height and weight | X | | | | | |
| Medical history, including COPD | X | | | | | |
| Concomitant treatment for COPD | X | | | | | |
| Routine laboratory blood tests | X[5] | | | | | |
| Vital signs | X | X | X | X | X | |
| COPD assessments | | | | | | |
| *Clinical assessment* | X | X | X | X | X | X |
| *Spirometry* | X | X | X | X | X | |
| *Blood gasses* | $X^2$ | $X^2$ | $X^2$ | $X^2$ | $X^2$ | |
| *Pulse oximetry ($SpO_2$)* | X | X | X | X | X | |
| *Respiration rate* | X | X | X | X | X | |
| N-Tidal C device | | | | | | |
| *Demonstration and training* | X | | | | | |
| *Use of device*[3] | X | X | X | X | $X^4$ | |
| *Check use of device* | X | X | X | X | X | |
| *Completion of patient diary* | X | X | X | X | X | |
| *Download capnometry data* | | X | X | X | X | |
| *Collect device* | | | | | X | |
| Adverse events | | X | X | X | X | X |
| Concomitant therapy | X | X | X | X | X | X |

Table 3: Schedule of assessments for COPD Breathing Record Study 2 (CBRS2).

| | **Screening** | **Monitoring Period** | | | | | **Follow-up** |
|---|---|---|---|---|---|---|---|
| | | **Week 0** | **Week 2** | **Week 10** | **Week 18** | **Week 26** | **Week 28** |
| Window (days) | -14 to 0 | | ±4 | ±4 | ±4 | ±3 | ±7 |
| **Study visit** | **1**[1] | **2**[1] | **3** | **4** | **5** | **6** | **7** |
| *Location* | *Clinic* | *Clinic* | *Clinic* | *Clinic* | *Clinic* | *Clinic* | *Telephone* |
| Informed consent | X | | | | | | |
| Demography | X | | | | | | |
| Height and weight | X | | | | | | |
| Medical history, including COPD | X | | | | | | |
| Physical exam - initial | X | | | | | | |
| Concomitant medicines review | X | X | X | X | X | X | |
| mMRC | X | | | | | | |
| CAT | X | | | | | | |
| Spirometry | X | | | | | | |
| COPD assessments | | | | | | | |
| *Vital Signs* | X | X | X | X | X | X | |
| *Respiration Rate* | X | X | X | X | X | X | |
| *Pulse oximetry ($SpO_2$)* | X | X | X | X | X | X | |
| *Physical exam - follow on* | | | | X | | X | |
| N-Tidal C device | | | | | | | |
| *Demonstration of device* | X | | | | | | |
| *Training on device* | | X | | | | | |
| *Use of device*[2] | | X | X | X | X | X[3] | |
| *Download N-Tidal C data* | | | X | X | X | X | |
| *Collect device* | | | | | | X | |
| *Explain patient diary* | X | X | | | | | |
| *Review and collect patient diary* | | | X | X | X | X | |
| *Supply new patient diary, batteries and mouthpieces* | | | X | X | X | | |
| *Device usage evaluation* | | | | | | | X |
| Adverse events | | X | X | X | X | X | X |
| Concomitant therapy | X | X | X | X | X | X | |

Table 4: Schedule of assessments for General Breathing Record Study (GBRS).

| **Condition / assessments** | **Baseline** | **2 months** | **4 months** | **6 months** | **At exacerbation** |
|---|---|---|---|---|---|
| **All Participants** | | | | | |
| Informed Consent | X | | | | |
| Demographics | X | | | | |
| Height & Weight | X | | | | |
| Medical History | X | | | | |
| Medication Review | X | X | X | X | X |
| Vital Signs | X | X | X | X | X |
| *N-Tidal C device* | | | | | |
| Demonstration and training | X | | | | |
| Check use of device | X | X | X | X | X |
| Download device data | | X | X | X | X |
| Collect device | | | | X | |
| **Asthma** | | | | | |
| Clinical Assessment | X | X | X | X | X |
| Symptom questionnaires (ACQ, AQLQ) and asthma symptom score | X | X | X | X | X |
| Spirometry | X | X | X | X | X |
| FeNO | X | X | X | X | X |
| PEFR | X | X | X | X | X |
| Peripheral Blood Eosinophil Count | X | | | | X |
| SPT (most recent within 3 years) | X | | | | |
| Full Body Plethysmography | X | | | X | |
| **Breathing pattern disorders** | | | | | |
| Clinical Assessment | X | | | | |
| Physiotherapist re-training & review | | X | X | X | |
| Diagnosis Questionnaires (Nijmegen, Pittsburgh Index) | X | | | | |
| Symptom Questionnaires (Nijmegen, Dyspnoea-12, VCDQ) | X | X | X | X | |
| Symptom diary review | X | X | X | X | X |
| Spirometry | X | | | | |
| Full Body Plethysmography | X | | | | |
| **Heart Failure** | | | | | |
| Clinical Assessment, weight, BP & HR | X | X | X | X | X |
| TTE | X | | | | X |
| NT pro-BNP | X | X | X | X | X |
| NYHA Class | X | X | X | X | X |
| Symptom questionnaire (KCCQ) | X | X | X | X | X |
| **Motor Neuron Disease** | | | | | |
| Clinical Assessment & Weight | X | X | X | X | X |
| Spirometry | X | X | X | X | |

Continued on next page

Table 4 – continued from previous page

| **Condition / assessments** | **Baseline** | **2 months** | **4 months** | **6 months** | **At exacerbation** |
|---|---|---|---|---|---|
| PEFR | X | X | X | X | X |
| Cough Assist Device data (if applicable) | X | X | X | X | X |
| Full Body Plethysmography (if possible) | (X) | | | (X) | |
| ABG | X | X | X | X | X |
| Ventilator data | X | X | X | X | X |
| Epworth Score | X | X | X | X | X |
| **Pneumonia (2 month cohort, extended to 4 months if unresolved)** | | | | | |
| Clinical assessment | X | X | (X) | | |
| CXR | X | X | (X) | | |
| FBC | X | | | | |
| CURB-65 Score | X | X | (X) | | |
| **Healthy volunteers** | | | | | |
| Clinical Assessment | X | | | X | |
| Spirometry | X | | | X | |
| Full Body Plethysmography | X | | | | |
| ECG | X | | | | |
| FeNO | X | | | X | |
| Symptom review – upper and lower respiratory tract symptoms | X | X | X | X | |

Table 5: Schedule of events for Asthma Breathing Record Study (ABRS).

| **ASSESSMENTS** | **Baseline** | **3 M** | **6 M** | **9 M*** | **12 M*** |
|---|---|---|---|---|---|
| **All Participants** | | | | | |
| Informed Consent | X | | | | |
| Demographics | X | | | | |
| Height & Weight | X | | | | |
| Medical History | X | | | | |
| Medication Review | X | X | X | X | X |
| Vital Signs | X | X | X | X | X |
| Message Dynamics PIS and consent form | X | | | | |
| Supply peak flow meter and check technique | X | | | | |
| Confirm and review personalised asthma action plan | X | X | X | X | X |
| *N-Tidal* | | | | | |
| Demonstration and training | X | | | | |
| Check use of device | X | X | X | X | X |
| Supply new mouthpieces and airways | X | | X | | |
| Collect device | | | X* | | X |

Table 6: Schedule of assessments for adult asthma in the Asthma Breathing Record Study (ABRS).

| **Adult Asthma ($\geq$ 16 years)** | **Baseline** | **3 M** | **6 M** | **9 M*** | **12 M*** |
|---|---|---|---|---|---|
| Clinical Assessment | X | X | X | X | X |
| Symptom questionnaires (ACQ, AQLQ) and asthma symptom score | X | X | X | X | X |
| Health related QoL questionnaires EQ 5D-5L | X | X | X* | X | X |
| WPAI | X | | X* | | |
| VAS questionnaire | | | X* | | X |
| Spirometry | X | X | X | X | X |
| *FeNO** | X | X | X | X | X |
| PEFR | X | X | X | X | X |
| *Most recent peripheral blood eosinophil count** | X | | | | |
| *SPT (most recent within 3 years)** | X | | | | |
| *Full Body Plethysmography** | X | | | | |
| *Oscillometry** | X | | X | | X |
| Health care resource use Questionnaire | X | X | X | X | X |

Table 7: Schedule of assessments for paediatric asthma in the Asthma Breathing Record Study (ABRS).

| **Paediatric Asthma (7-15 years)** | **Baseline** | **3 M** | **6 M** | **9 M** | **12 M** |
|---|---|---|---|---|---|
| Clinical Assessment | X | X | X | | |
| Symptom questionnaires (ACQ, PAQLQ) and asthma symptom score | X | X | X | | |
| VAS questionnaire | | | X | | |
| *Spirometry** | X | X | X | | |
| *FeNO** | X | X | X | | |
| PEFR | X | X | X | | |
| *Most recent peripheral Blood Eosinophil Count** | X | | | | |
| *SPT (most recent within 3 years) ** | X | | | | |
| *Oscillometry ** | X | | X | | |
| *Health care resource use Questionnaire* | | X | X | | |
